# Epidemiology of *Burkholderia pseudomallei, Streptococcus suis, Salmonella* spp., *Shigella* spp. and *Vibrio* spp. infections in 111 hospitals in Thailand, 2022

**DOI:** 10.1101/2024.05.30.24307969

**Authors:** Charuttaporn Jitpeera, Somkid Kripattanapong, Preeyarach Klaytong, Chalida Rangsiwutisak, Prapass Wannapinij, Pawinee Doungngern, Papassorn Pinyopornpanish, Panida Chamawan, Voranadda Srisuphan, Krittiya Tuamsuwan, Phairam Boonyarit, Orapan Sripichai, Soawapak Hinjoy, John Stelling, Paul Turner, Wichan Bhunyakitikorn, Sopon Iamsirithaworn, Direk Limmathurotsakul

**Affiliations:** Division of Epidemiology, Department of Disease Control, Ministry of Public Health, Nonthaburi 11000, Thailand; Mahidol-Oxford Tropical Medicine Research Unit, Faculty of Tropical Medicine, Mahidol University, Bangkok, 10400, Thailand; Division of Communicable Diseases, Department of Disease Control, Ministry of Public Health, Nonthaburi, 11000, Thailand; Health Administration Division, The Office of Permanent Secretary, Ministry of Public Health, Nonthaburi, 11000, Thailand; Department of Medical Science, Ministry of Public Health, Nonthaburi, 11000, Thailand (OS); Office of International Cooperation, Department of Disease Control, Ministry of Public Health, Nonthaburi 11000, Thailand (SH); Brigham and Women’s Hospital and Harvard Medical School, Boston, MA, United States (JS); Cambodia-Oxford Medical Research Unit, Angkor Hospital for Children, Siem Reap, Cambodia; Centre for Tropical Medicine and Global Health, University of Oxford, Oxford, United Kingdom; Department of Disease Control, Ministry of Public Health, Nonthaburi 11000, Thailand (SI); Department of Tropical Hygiene, Faculty of Tropical Medicine, Mahidol University, Bangkok, 10400, Thailand

**Keywords:** melioidosis, salmonella, shigella, *Streptococcus suis*, vibrio, incidence, mortality

## Abstract

The information on notifiable diseases in low- and middle-income countries is often incomplete, limiting our understanding of their epidemiology. Our study addresses this knowledge gap by analyzing microbiology laboratory and hospital admission data from 111 of 127 public referral hospitals in Thailand, excluding Bangkok, from January to December 2022. We evaluated factors associated with the incidence of notifiable bacterial diseases (NBDs) caused by 11 pathogens; including *Brucella* spp., *Burkholderia pseudomallei*, *Corynebacterium diphtheriae*, *Neisseria gonorrhoeae*, *Neisseria meningitidis*, non-typhoidal *Salmonella* spp. (NTS), *Salmonella enterica* serovar Paratyphi, *Salmonella enterica* serovar Typhi, *Shigella* spp., *Streptococcus suis*, and *Vibrio* spp.. We used multivariable Poisson random-effects regression models. Additionally, we compared their yearly incidence rates in 2022 with those from 2012-2015 in hospitals where paired data were available. In 2022, the NBD associated with the highest total number of deaths was *B. pseudomallei* (4,407 patients; 1,219 deaths) infection, followed by NTS (4,501 patients; 461 deaths) and *S. suis* (867 patients, 134 deaths) infection. The incidence rate of *B. pseudomallei* and *S. suis* infection was highest in the northeast and upper central, respectively. The incidence rate of NTS infection was not associated with geographical region. The yearly incidence rate of *B. pseudomallei* and *S. suis* infection in 2022 were higher than those between 2012-2015, while those of fecal-oral transmitted NBDs including NTS infection, typhoid, shigellosis and vibriosis were lower. Overall, *B. pseudomallei* and *S. suis* infection are emerging and associated with a high number of deaths in Thailand. Specific public health interventions are warranted.

## Introduction

Timely, reliable and complete information regarding notifiable diseases is essential for disease control and prevention, and enhancing our understanding of their epidemiology [1–3]. To achieve timeliness and completeness in data reporting, many high-income and upper-middle-income countries have strengthened their national surveillance systems by modernizing tools, technology and strategies [4–6]. These include automatic electronic laboratory-based reporting of notifiable diseases [5–7]. However, most low and middle-income countries (LMICs) have a shortage of resources, still use conventional or semi-automated data reporting systems, and do not automatically integrate laboratory data into their surveillance systems [4,8]. Therefore, the information available in the national surveillance systems of LMICs are still largely incomplete [9–11] and our understanding of their epidemiology remains limited.

In Thailand, the national surveillance systems monitors 13 dangerous communicable diseases and 57 notifiable diseases, overseen by the Department of Disease Control (DDC), Ministry of Public Health (MoPH) [12]. For the 13 dangerous communicable disease (e.g. Ebola and smallpox), immediate reporting of any suspected cases is required. For the 57 notifiable diseases, the reporting systems can be semi-automatic, utilizing the electronic data of final diagnosis based on the International Classification of Diseases, 10^th^ revision (ICD-10) recorded in the hospital information systems (HIS). However, the ICD-10 is reliable only in few conditions [13,14] and the surveillance systems do not utilize microbiology laboratory data. Recent studies showed that many public hospitals, which have microbiology laboratories, do not report most cases and deaths following culture-confirmed notifiable bacterial diseases (NBDs) to the national surveillance systems [11,15]. The incomplete data hinders efforts to improve awareness, control, prevention, and our understanding of NBDs in the country [16].

To overcome limitations in LMICs, we developed the AMASS (AutoMated tool for Antimicrobial resistance Surveillance System), an offline application that enables hospitals to automatically analyse and generate standardized antimicrobial resistance (AMR) surveillance reports from routine microbiology and hospital data [17]. The AMASS version 1.0 was released on 1^st^ February 2019 and tested in seven hospitals in seven countries [17]. We conceptualized that the AMASS could additionally analyse and generate summary reports for multiple NBDs [18]. The AMASS version 2.0 (AMASSv2.0) was released on 16^th^ May 2022 and tested using data of 49 public hospitals in Thailand from 2012 to 2015 [15]. We demonstrate that national statistics on NBDs in LMICs could be improved by integrating information from readily available databases [15]. In 2023, collaborating with Health Administration Division, MoPH Thailand, we implemented AMASSv2.0 in 127 public hospitals in Thailand using the data from 2022 [19]. We recently reported the epidemiology of AMR bloodstream infections in 111 hospitals [20].

Here we aimed to evaluate the epidemiology of multiple NBDs in 111 hospitals in Thailand using the data from 2022. We also examined the trends of each NBD by comparing the data from 2022 with the data from 2012 to 2015.

## Methods

### Study setting

In 2022, Thailand had a population of 66.1 million, consisted of 77 provinces, and covered 513,120 km^2^. The health systems in each province were integrated into 12 groups of provinces, known as health regions, plus the capital Bangkok as health region 13 (Figure 1), using the concept of decentralization [21]. The Health Administration Division, Ministry of Public Health (MoPH) Thailand, supervised 127 public referral hospitals in health regions 1 to 12. These included 35 advanced-level referral hospital (i.e. level A, with a bed size of about 500-1,200 beds), 55 standard-level referral hospital (i.e. level S, with a bed size of about 300 to 500) and 37 mid-level referral hospital (i.e. level M1, with a bed size of about 180-300) [22]. All level A and S hospitals, and most of level M1 hospitals were equipped with a microbiology laboratory capable of performing bacterial culture using standard methodologies for bacterial identification and susceptibility testing provided by the Department of Medical Sciences, MoPH, Thailand [23].

**Figure 1.**
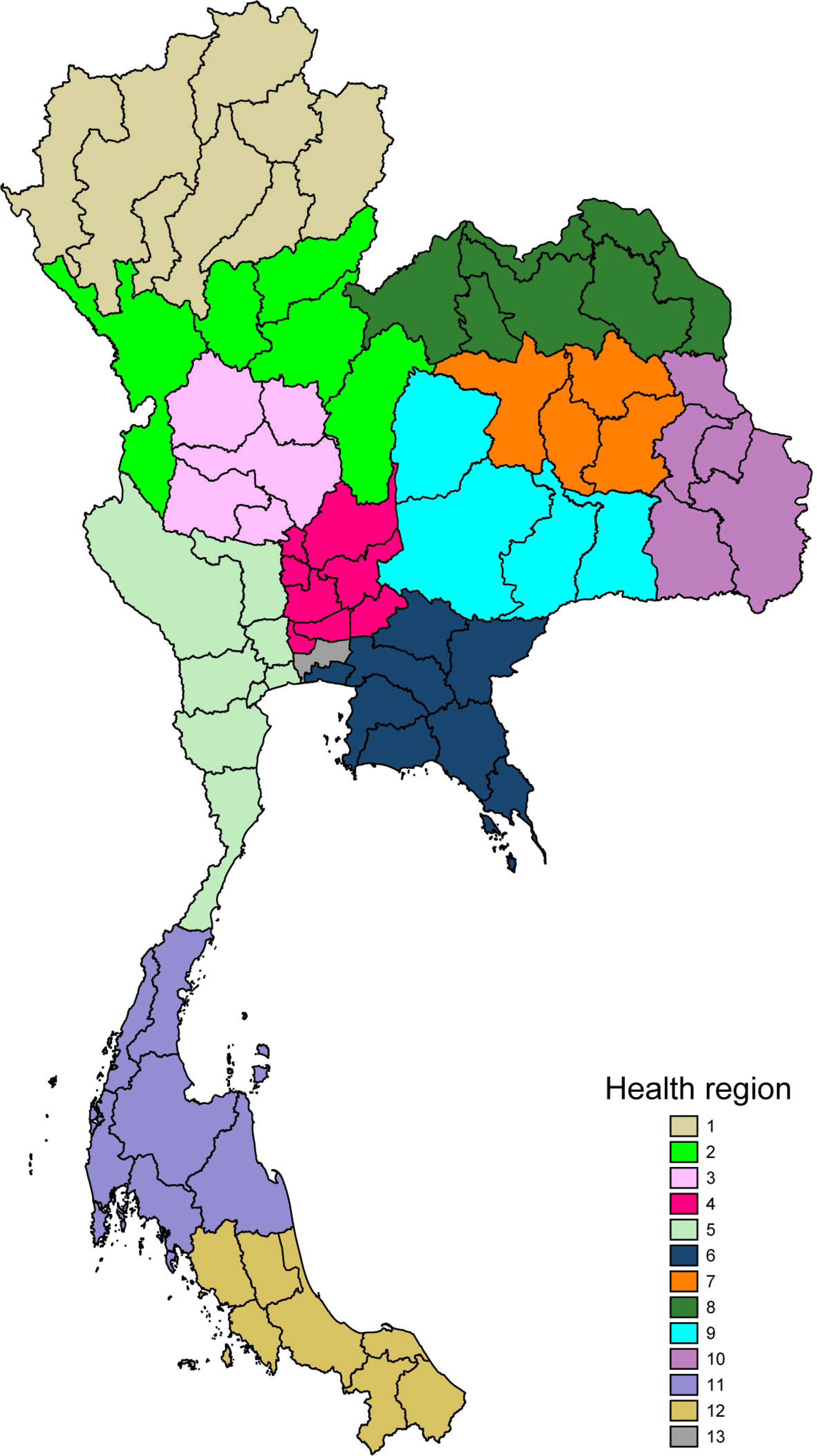
Health regions in Thailand **Footnote of figure 1.** National Health Security Office (NHSO) Region 1 is in Chiang Mai, 2 in Phitsanulok, 3 in Nakhonsawan, 4 in Saraburi, 5 in Ratchaburi, 6 in Rayong, 7 in Khon Kaen, 8 in Udon Thani, 9 in Nakonratchasima, 10 in Ubonratchathani, 11 in Suratthani, 12 in Songkhla and 13 in Bangkok.

From 16 December 2022 to 30 June 2023, on behalf of the Health Administration Division, MoPH, we invited and trained 127 public referral hospitals in health regions 1 to 12 to utilize the AMASS with their own microbiology and hospital admission data files via four online meetings, five face-to-face meetings and on-line support [20]. Subsequently, the hospitals that completed utilization of the AMASS submitted summary data generated by the tool to the MoPH [20].

### Study design

We conducted a retrospective study evaluating epidemiology of selected NBDs diagnosed by culture using microbiology laboratory and hospital admission data from 2022. We also compared the yearly incidence rates of each NBD in 2022 with those from 2012-2015, using paired data from 49 public referral hospitals that were previously published [15].

The NBDs under evaluation included infections caused by 12 pathogens; *Brucella* spp., *Burkholderia pseudomallei*, *Corynebacterium diphtheriae*, *Neisseria gonorrhoeae, Neisseria meningitidis,* Non-typhoidal *Salmonella* spp., *Salmonella enterica* serovar Paratyphi, *Salmonella enterica* serovar Typhi, *Shigella* spp., *Streptococcus suis*, and *Vibrio* spp. infections. NBD cases were defined as having any clinical specimen (including blood, respiratory tract specimens, urine, stool, cerebrospinal fluid, genital swabs and other specimens) culture positive for a pathogen. The AMASS merged microbiology laboratory and hospital admission data using the hospital number (i.e. the patient identifier) present in both data files. Then, the AMASS deduplicated the data reporting total number of inpatients with a clinical specimen culture positive for a pathogen during the evaluation period.^11^ Mortality was defined using the discharge summary (in the hospital admission data) which was routinely completed by the attending physician and reported to the MoPH. For each NBD, in case that a patient was admitted with that NBD more than once during the evaluation period, the mortality outcome of the first admission was presented.

### Statistical Analysis

Data were summarized with medians and interquartile ranges (IQR) for continuous measures, and proportions for discrete measures. IQRs are presented in terms of 25th and 75th percentiles. Continuous variables and proportions were compared between groups using Kruskal Wallis tests and Chi-square tests, respectively.

For NBDs with more than 100 cases in the year 2022, we evaluated factors associated with the incidence rate of NBDs per 100,000 population using multivariable Poisson random-effects regression models [24]. The total number of NBD cases in each province was calculated by summing the number of NBD cases from all hospitals located in the same province. We assumed that the distribution of province-specific random effects was normal. Factors evaluated included health region, Gross Provincial Product (GPP), pig density and poultry density. Data of GPP in 2021 [25] were used as a proxy for the size of the economy in each province. Pig density and poultry density (per square meters) were estimated by using the total number of pigs and poultry in each province in Thailand in 2022, divided by the total area of each province [26].

Additionally, we compared the yearly incidence rates of each NBD in 2022 with the those between 2012-2015 in hospitals where paired data were available. Multivariable Poisson random-effects regression models were used to evaluate the change of incidence rate per 100,000 population between the time periods. We also compared the total number of cases and deaths of each NBD diagnosed by culture in 2022 in our study with those of relevant notifiable diseases reported to the NNDSS of Thailand [27]. We used STATA (version 14.2; College Station, Texas) for all analyses (Appendix A).

### Data availability

The hospital-level summary data used for the study are open-access and available at https://figshare.com/s/79f2b4b9422263b9048b.

### Ethics

Ethical permission for this study was obtained from the Institute for the Committee of the Faculty of Tropical Medicine, Mahidol University (TMEC 23-085). Individual patient consent was not sought as this was a retrospective study, and the Ethical and Scientific Review Committees approved the process.

## Results

### Baseline characteristics

Of 127 public referral hospitals, 116 (91%) used the AMASS to analyze their microbiology and hospital admission data files, and submitted the summary AMR and NBD data from 2022 to the MoPH. Four hospitals had incomplete microbiology data, and one hospital had incomplete hospital admission data. These hospitals were removed from the analysis. Therefore, a total of 111 hospitals were included in the final analysis.

Of all public referral hospitals in Thailand, 100% of Level A hospitals (35/35), 89% of Level S hospitals (49/55) and 73% of Level M1 hospitals (27/37) were included in this study. Data were available from 74 of 77 provinces (96%) in Thailand, all provinces except Mae Hong Son, Nakorn Nayok and Bangkok.

A total of 46 hospitals in 42 provinces had paired data from 2022 and 2012-2015.

### Brucella spp

In 2022, there were 11 cases with culture-confirmed *Brucella* spp. infection (Figure 2A) and 1 of them died (in-hospital mortality 9%). Among provinces where paired data were available, the incidence rate did not differ between the time periods (p=0.15).

**Figure 2.**
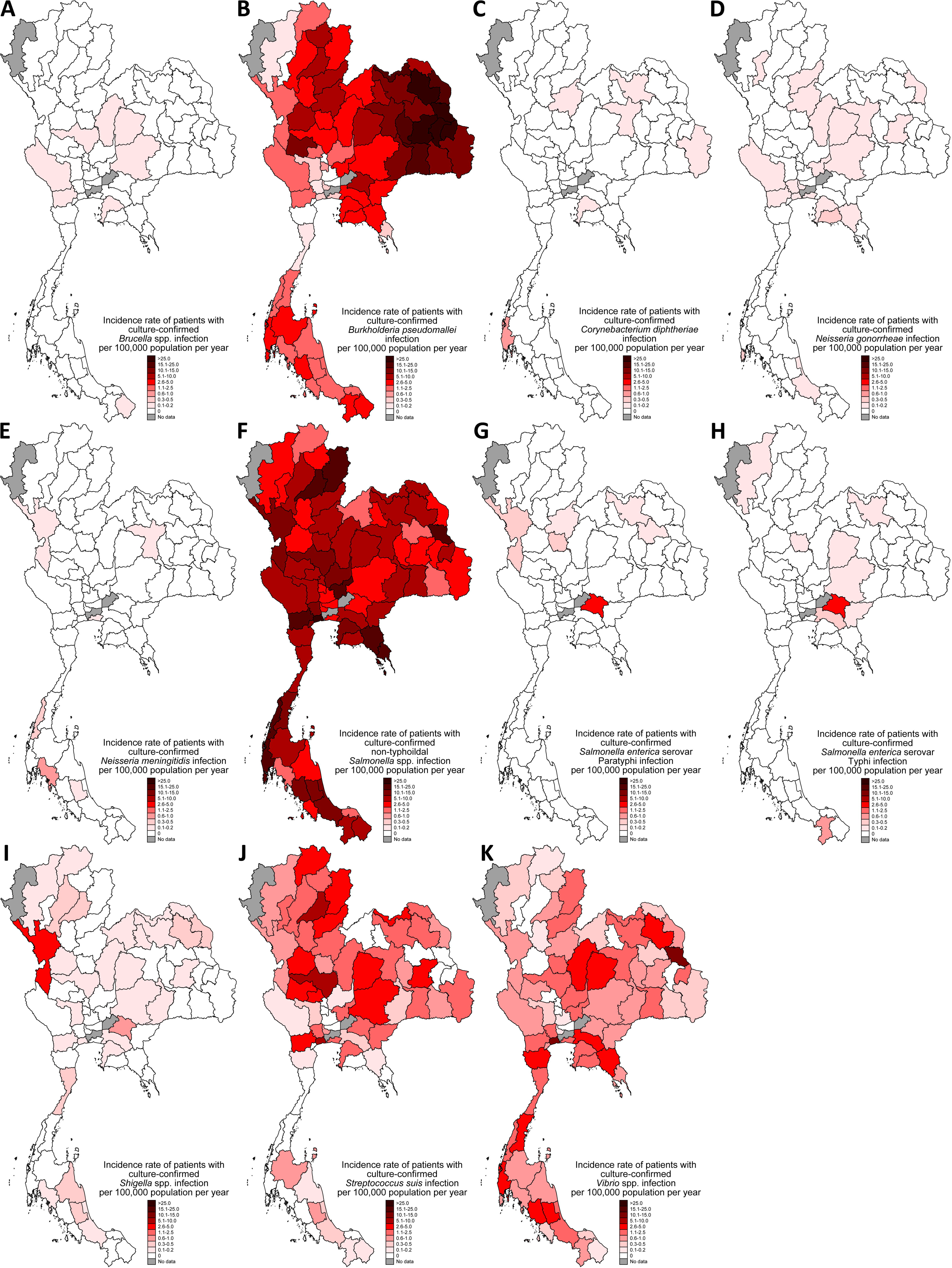
Incidence rate of cases with notifiable bacterial diseases diagnosed by culture per 100,000 population in 2022 in Thailand

### B. pseudomallei

In 2022, there were 4,407 cases with culture-confirmed *B. pseudomallei* infection and 1,219 of them died (in-hospital mortality 27.7%). The incidence rate (Figure 2B) was highest in the northeast (health regions 7, 8, 9 and 10), followed by the upper central (health region 3), north (health region 1 and 2) east (health region 6) and south (health region 11 and 12). The incidence rate was lowest in the lower central (health region 4) and west (health region 5). In the multivariable models, health region and poultry density (adjust incidence rate ratio [aIRR] 1.31, 95%CI 1.02-1.69, p=0.04) were independently associated with the incidence of *B. pseudomallei* infection per 100,000 population (Table S1). GPP and pig density and poultry density were not independently associated with the incidence rate (both p>0.20)

Among provinces where paired data were available, the yearly incidence rate of *B. pseudomallei* infection in 2022 was higher than that between 2012-2015 by 58% (aIRR 1.58, 95%CI 1.49-1.68, p<0.001).

### C. diphtheriae

In 2022, there were 10 cases with culture-confirmed *C. diphtheriae* infection (Figure 2C) and 1 of them died (in-hospital mortality 10%). Among provinces where paired data were available, the incidence rate did not differ between the time periods (p=0.29).

### N. gonorrhoeae

In 2022, there were 25 cases with culture-confirmed *N. gonorrhoeae* infection (Figure 2D) and none died. Among provinces where paired data were available, the incidence rate did not differ between the time periods (p=0.33).

### N. meningitidis

In 2022, there were 9 cases with culture-confirmed *N. meningitidis* infection (Figure 2E) and 2 died (in-hospital mortality 22%). Among provinces where paired data were available, the incidence rate did not differ between the time periods (p=0.55).

### Non-typhoidal *Salmonella* spp. (NTS)

In 2022, there were 4,501 cases with culture-confirmed NTS infection (Figure 2F) and 461 died (in-hospital mortality 10.2%). In the multivariable models, health region, GPP, pig density and poultry density were not associated with the incidence rate (all p>0.20, Table S2).

Among provinces where paired data were available, the yearly incidence rate of NTS cases in 2022 was lower than that between 2012-2015 by 37% (aIRR 0.63, 95%CI 0.60-0.67, p<0.001).

### Salmonella enterica serovar Paratyphi

In 2022, there were 30 cases with culture-confirmed *Salmonella enterica* serovar Paratyphi infection (Figure 2G) and 4 died (in-hospital mortality 13%). Among provinces where paired data were available, the incidence rate did not differ between the time periods (p=0.39).

### Salmonella enterica serovar Typhi

In 2022, there were 32 cases with culture-confirmed *Salmonella enterica* serovar Typhi infection (Figure 2H) and 6 died (in-hospital mortality 19%).

Among provinces where paired data were available, the yearly incidence rate of *Salmonella enterica* serovar Typhi infection in 2022 was lower than that between 2012-2015 by 83% (aIRR 0.17, 95%CI 0.07-0.41, p<0.001).

### Shigella spp

In 2022, there were 68 cases with culture-confirmed *Shigella* spp. infection (Figure 2I) and 4 died (in-hospital mortality 6%).

Among provinces where paired data were available, the yearly incidence rate of *Shigella* spp. infection in 2022 was lower than that between 2012-2015 by 78% (aIRR 0.22, 95%CI 0.14-0.36, p<0.001).

### S. suis

In 2022, there were 867 cases with culture-confirmed *S. suis* infection and 134 of them died (in-hospital mortality 15.5%). The incidence rate (Figure 2J) was highest in the upper central (health regions 3), followed by the north (health regions 1 and 2) and northeast (health regions 7, 8, 9 and 10). The incidence rate was lowest in the south (health regions 11 and 12) and lower central (health region 4). In the multivariable models, health region was associated with the incidence of *S. suis* infection per 100,000 population (Table S3). GPP, pig density and poultry density were not independently associated with the incidence rate.

Among provinces where paired data were available, the yearly incidence rate of *S. suis* infection in 2022 was higher than that between 2012-2015 by 172% (aIRR 2.72, 95%CI 2.29-3.24, p<0.001).

### Vibrio spp

In 2022, there were 809 cases with culture-confirmed *Vibrio* spp. infection and 122 of them died (in-hospital mortality 15.1%). The crude incidence rate (Figure 2K) was highest in the west (health region 5), followed by the east (health region 6). The crude incidence rate was lowest in the central (health region 3 and 4) and upper north (health region 1). In the multivariable models, health region was associated with the incidence of *Vibrio* spp. infection per 100,000 population (Table S4). GPP, pig density and poultry density were not independently associated with the incidence rate.

Among provinces where paired data were available, the yearly incidence rate of *Vibrio* spp. infection in 2022 was lower than that between 2012-2015 by 25% (aIRR 0.75, 95%CI 0.66-0.85, p<0.001).

### Comparison with the national surveillance systems

In 2022, the total number of cases (4,407 vs. 3,573 cases) and deaths (1,219 vs. 157 deaths) following *B. pseudomallei* infection diagnosed by culture was higher than those reported to the national surveillance systems (Table 1). The total number of cases (867 vs. 383 cases) and deaths (134 vs. 10 deaths) following *S. suis* infection diagnosed by culture was also higher. The total number of deaths following fecal-oral transmitted NBDs diagnosed by culture (including 461 for non-typhoidal salmonella, 122 for vibriosis, 4 for paratyphoid, 6 for typhoid and 4 for shigellosis) were also higher than those reported to the national surveillance systems (0 deaths for all relevant fecal-oral transmitted NBDs).

**Table 1.** Total number of cases and deaths following selected notifiable bacterial diseases (NBDs) diagnosed by culture in 111 hospitals) compared with total number of cases and deaths with relevant notifiable diseases reported to the national surveillance systems (NSS) in Thailand in 2022.

| **Total number of cases \*** | **This study** | **Relevant notifiable diseases in the NSS** | **NSS** |
| --- | --- | --- | --- |
| *Brucella* spp. infection | 11 | Brucellosis | 15 |
| *Burkholderia pseudomallei* infection | 4,407 | Melioidosis | 3,573 |
| *Corynebacterium diphtheriae* infection | 10 | Diphtheria | 0 |
| *Neisseria gonorrhoeae* infection | 25 | Gonorrhea | 6,915 |
| *Neisseria meningitidis* infection | 9 | Meningococcal meningitis | 19 |
| Non-typhoidal *Salmonella* spp. and *Vibrio* spp. infection | 4,501 and 809 | Food poisoning and cholera | 72,439 and 4 |
| *Salmonella enterica* serovar Paratyphi infection | 30 | Paratyphoid | 91 |
| *Salmonella enterica* serovar Typhi infection | 32 | Typhoid | 754 |
| *Shigella* spp. infection | 68 | Bacillary dysentery | 377 |
| *Streptococcus suis* infection | 867 | *Streptococcus suis* infection | 383 |
| **Total number of deaths** |  |  |  |
| *Brucella* spp. infection | 1 | Brucellosis | 0 |
| *Burkholderia pseudomallei* infection | 1,219 | Melioidosis | 157 |
| *Corynebacterium diphtheriae* infection | 1 | Diphtheria | 0 |
| *Neisseria gonorrhoeae* infection | 0 | Gonorrhea | 0 |
| *Neisseria meningitidis* infection | 2 | Meningococcal meningitis | 3 |
| Non-typhoidal *Salmonella* spp. and *Vibrio* spp. infection | 461 and 122 | Food poisoning and cholera | 0 and 0 |
| *Salmonella enterica* serovar Paratyphi infection | 4 | Paratyphoid | 0 |
| *Salmonella enterica* serovar Typhi infection | 6 | Typhoid | 0 |
| *Shigella* spp. infection | 4 | Bacillary dysentery | 0 |
| *Streptococcus suis* infection | 134 | *Streptococcus suis* infection | 10 |

\* Case was defined as an inpatient with a clinical specimen culture positive for a pathogen during the evaluation period, while the NSS included reports of suspected, probable and confirmed cases using a wide range of case definitions of each notifiable disease (Appendix B).

## Discussion

Our findings provide evidence that the incidences of melioidosis and *S. suis* infection are increasing and associated with a high number of deaths in Thailand in 2022. We also show that fecal-oral transmitted NBDs including non-typhoidal salmonellosis, typhoid, shigellosis and vibriosis are still present and associated with deaths, but their incidence rates are decreasing compared to the data from 2012-2015. This study highlights the potential advantage of utilization of routine microbiology and hospital admission data from hospitals. The local and timely data of NBDs can supplement and monitor the performance of the national surveillance systems. The accurate data can consequently identify diseases and areas with high burden, improve public health interventions, and prioritize resource allocation.

The finding that more than 1,200 deaths following melioidosis in 2022 is alarming. This finding is consistent with a previous modelling study predicting that the total number of deaths following melioidosis could range from 1,259 to 6,678 in Thailand if all patients were tested with bacterial culture and data were reported nationwide [28]. The increased incidence rate of melioidosis could be associated with the increasing incidence of diabetes (the major risk factor of melioidosis) and improvement of diagnostic stewardship (i.e. utilization of culture) and bacterial identification in public hospitals in Thailand over time [16,29,30]. The association between poultry density and melioidosis is unclear, and could be a spurious finding. Further studies and actions to reduce the burden of melioidosis in Thailand are urgently needed [31].

Similarly, the increase in *S. suis* infection is alarming. This could be associated with an increase in consumption of undercooked pork products [32–34] and an increase in infected meat in the market [35]. The latter is a concern following the news of the large illegal pork imported to Thailand since 2021 after the shortage of domestic pork due to the outbreak of African swine fever in Thailand [36]. The DDC will utilize the data to additionally implement and enforce behavioral interventions, education and food biosafety in the country [34].

The decrease of multiple fecal-oral transmitted diseases could be due to improvement in clean water supply, sanitation and related health intervention programs over time [37,38]. The MoPH and related stakeholders should maintain and strengthen the public health interventions to decrease incidences of these infections further.

Our study and approach have several strengths. First, our study included most of the public referral hospitals in the country. Second, we utilize microbiology laboratory data, which is highly specific to the diagnosis of NBDs. Third, although our approach is semi-automatic, this approach is easy to scale up in LMICs because the AMASS programme is open-access, highly-compatible, and user-friendly without the need for data experts with adequate skills in statistical software [17,19].

Our study and approach have several limitations. First, our approach included only inpatients in the public referral hospitals. Therefore, our estimates did not include patients who did not require hospitalization, and those who were hospitalized in private, military or university hospitals. Nonetheless, majority of healthcare services in Thailand are in the public sector [39]. Second, our approach is not applicable in settings where microbiology and hospital admission data are not computerized. Third, our approach focused on bacterial culture results. Therefore, the findings could be influenced by diagnostic stewardship and, capability and expertise of the microbiology laboratories. Fourth, our approach utilizes summary data and could not evaluate individual-level clinical data and microbiology laboratory data in details. Fifth, the in-hospital mortality could be lower than the all-cause mortality because a preference to die at home is high in some regions in Thailand [11].

In conclusion, the burden of melioidosis and *S. suis* infection are increasing in Thailand. Their incidence rates are higher in some regions than in others. Specific public health interventions to reduce the burden of melioidosis and *S. suis* infection are urgently required.

## Supporting information

Suppementary file

## Acknowledgement

We gratefully acknowledge the laboratory team and IT team of all hospitals for their participation and support.

## Declaration of interest statement

The authors report there are no competing interests to declare.

## Funding

This research was supported by the Wellcome Trust under Grant [number 224681/Z/21/Z]. For the purpose of Open Access, the author has applied a CC BY public copyright licence to any Author Accepted Manuscript version arising from this submission.

## Notes

### Competing Interest Statement

The authors have declared no competing interest.

### Funding Statement

This research was funded in part by the Wellcome Trust [224681/Z/21/Z]. For the purpose of Open Access, the author has applied a CC BY public copyright licence to any Author Accepted Manuscript version arising from this submission.

### Summary of Updates

The analysis has been revised, and the text have been updated accordingly.

