## Supplementary material for "Epidemiology of *Burkholderia pseudomallei, Streptococcus suis, Salmonella* spp., *Shigella* spp. and *Vibrio* spp. infections in 111 hospitals in Thailand, 2022": Suppementary file

- **Table S1. Factors associated with the incidence of cases with culture-confirmed *Burkholderia pseudomallei* infection per 100,000 population in 74 provinces in Thailand, 2022**

| **Factors*** | **No. of cases** | **Population** | **Crude incidence rate ratio (95% CI)** | **P value** | **Adjusted incidence rate ratio (95% CI)** | **P value** |
| --- | --- | --- | --- | --- | --- | --- |
| Health regions |  |  |  |  |  |  |
| 1 | 136 | 5,725,000 | 4.0 (1.6-10.2) | <0.001 | 7.4 (2.6-21.2) | <0.001 |
| 2 | 178 | 3,723,000 | 8.0 (3.0-21.5) |  | 14.7 (4.9-44.0) |  |
| 3 | 163 | 2,996,000 | 9.5 (3.6-25.6) |  | 14.9 (5.1-43.2) |  |
| 4 | 46 | 5,275,000 | 1.0 |  | 1.0 |  |
| 5 | 38 | 5,430,000 | 0.8 (0.3-2.2) |  | 1.1 (0.3-3.2) |  |
| 6 | 213 | 6,312,000 | 5.4 (2.2-13.2) |  | 6.5 (2.4-17.5) |  |
| 7 | 913 | 5,131,000 | 33.1 (11.9-92.6) |  | 57.4 (18.7-176.5) |  |
| 8 | 905 | 5,629,000 | 22.0 (8.8-54.5) |  | 39.6 (14.0-112.1) |  |
| 9 | 682 | 6,924,000 | 18.2 (6.5-50.8) |  | 29.7 (9.8-89.7) |  |
| 10 | 910 | 4,702,000 | 46.3 (17.5-122.6) |  | 83.2 (28.0-247.2) |  |
| 11 | 103 | 4,561,000 | 4.1 (1.6-10.4) |  | 7.3 (2.5-20.9) |  |
| 12 | 120 | 5,084,000 | 4.3 (1.7-10.9) |  | 7.2 (2.6-20.2) |  |
| Gross provincial product* | - | - | 0.68 (0.49-0.95) | 0.024 | 1.01 (0.78-1.30) | 0.97 |
| Pig density* | - | - | 0.88 (0.62-1.23) | 0.45 | 1.10 (0.87-1.38) | 0.42 |
| Poultry density* | - | - | 0.82 (0.59-1.16) | 0.27 | 1.31 (1.02-1.69) | 0.04 |

- * Data were from 111 public referral hospitals in Thailand. All three continuous variables were standardized by centering at the mean values and divided by the standard deviation. The multivariable model consists of 4,407 patients with *Burkholderia pseudomallei* infection from 61,492,000 population over one year in 2022.
- **Table S2. Factors associated with the incidence of cases with culture-confirmed non-typhoidal *Salmonella* spp. (NTS) infection per 100,000 population in 74 provinces in Thailand, 2022**

| **Factors*** | **No. of cases** | **Population** | **Crude incidence rate ratio (95% CI)** | **P value** | **Adjusted incidence rate ratio (95% CI)** | **P value** |
| --- | --- | --- | --- | --- | --- | --- |
| Health regions |  |  |  |  |  |  |
| 1 | 391 | 5,725,000 | 2.0 (1.0-4.4) | 0.14 | 2.1 (1.0-4.4) | 0.47 |
| 2 | 283 | 3,723,000 | 2.3 (1.0-5.2) |  | 2.4 (1.1-5.2) |  |
| 3 | 233 | 2,996,000 | 2.2 (1.0-4.9) |  | 2.1 (0.9-4.6) |  |
| 4 | 409 | 5,275,000 | 2.1 (1.0-4.5) |  | 1.8 (0.8-4.0) |  |
| 5 | 620 | 5,430,000 | 3.1 (1.5-6.5) |  | 2.8 (1.3-5.9) |  |
| 6 | 584 | 6,312,000 | 2.8 (1.3-5.8) |  | 2.3 (1.0-5.4) |  |
| 7 | 212 | 5,131,000 | 1.0 |  | 1.0 |  |
| 8 | 299 | 5,629,000 | 1.4 (0.7-3.1) |  | 1.5 (0.7-3.1) |  |
| 9 | 429 | 6,924,000 | 2.0 (0.9-4.7) |  | 2.0 (0.9-4.6) |  |
| 10 | 220 | 4,702,000 | 1.7 (0.7-3.7) |  | 1.7 (0.8-3.8) |  |
| 11 | 348 | 4,561,000 | 2.8 (1.3-5.9) |  | 2.7 (1.3-5.8) |  |
| 12 | 473 | 5,084,000 | 2.3 (1.1-4.8) |  | 2.2 (1.1-4.7) |  |
| Gross provincial product* | - | - | 1.16 (1.00-1.35) | 0.047 | 1.05 (0.86-1.27) | 0.63 |
| Pig density* | - | - | 1.12 (0.97-1.30) | 0.13 | 1.03 (0.87-1.22) | 0.71 |
| Poultry density* | - | - | 1.12 (0.97-1.31) | 0.13 | 1.08 (0.89-1.30) | 0.44 |

- * Data were from 111 public referral hospitals in Thailand. All three continuous variables were standardized by centering at the mean values and divided by the standard deviation. The multivariable model consists of 4,501 patients with NTS infection from 61,492,000 population over one year in 2022.
- **Table S3. Factors associated with the incidence of cases with culture-confirmed *S. suis* infection per 100,000 population in 74 provinces in Thailand, 2022**

| **Factors*** | **No. of cases** | **Population** | **Crude incidence rate ratio (95% CI)** | **P value** | **Adjusted incidence rate ratio (95% CI)** | **P value** |
| --- | --- | --- | --- | --- | --- | --- |
| Health regions |  |  |  |  |  |  |
| 1 | 121 | 5,725,000 | 23.0 (5.1-104.8) | <0.001 | 23.0 (5.1-102.4) | <0.001 |
| 2 | 81 | 3,723,000 | 22.2 (4.5-109.8) |  | 22.2 (4.6-108.0) |  |
| 3 | 119 | 2,996,000 | 33.3 (6.7-164.4) |  | 27.5 (5.7-133.1) |  |
| 4 | 22 | 5,275,000 | 2.0 (0.4-10.1) |  | 1.5 (0.3-8.2) |  |
| 5 | 105 | 5,430,000 | 7.9 (1.7-35.7) |  | 4.9 (1.1-22.7) |  |
| 6 | 59 | 6,312,000 | 5.8 (1.3-26.4) |  | 4.4 (0.9-22.8) |  |
| 7 | 77 | 5,131,000 | 9.1 (1.7-49.0) |  | 8.6 (1.6-45.7) |  |
| 8 | 69 | 5,629,000 | 8.4 (1.8-38.6) |  | 8.1 (1.8-37.4) |  |
| 9 | 133 | 6,924,000 | 15.7 (3.0-82.8) |  | 14.3 (2.7-74.6) |  |
| 10 | 57 | 4,702,000 | 3.5 (0.6-19.4) |  | 3.6 (0.7-19.7) |  |
| 11 | 10 | 4,561,000 | 1.0 |  | 1.0 |  |
| 12 | 14 | 5,084,000 | 1.8 (0.4-9.3) |  | 1.6 (0.3-8.0) |  |
| Gross provincial product* | - | - | 0.77 (0.51-1.17) | 0.22 | 0.97 (0.62-1.51) | 0.89 |
| Pig density* | - | - | 1.28 (0.90-1.84) | 0.17 | 1.28 (0.94-1.74) | 0.11 |
| Poultry density* | - | - | 1.04 (0.71-1.51) | 0.84 | 1.16 (0.80-1.68) | 0.43 |

- * Data were from 111 public referral hospitals in Thailand. All three continuous variables were standardized by centering at the mean values and divided by the standard deviation. The multivariable model consists of 867 patients with *S. suis* infection from 61,492,000 population over one year in 2022.
- **Table S4. Factors associated with the incidence of cases with culture-confirmed *Vibrio* spp. infection per 100,000 population in 74 provinces in Thailand, 2022**

| **Factors*** | **No. of cases** | **Population** | **Crude incidence rate ratio (95% CI)** | **P value** | **Adjusted incidence rate ratio (95% CI)** | **P value** |
| --- | --- | --- | --- | --- | --- | --- |
| Health regions |  |  |  |  |  |  |
| 1 | 23 | 5,725,000 | 1.1 (0.3-3.9) | 0.033 | 2.2 (0.5-9.0) | 0.032 |
| 2 | 51 | 3,723,000 | 3.5 (1.0-12.5) |  | 6.8 (1.6-28.6) |  |
| 3 | 20 | 2,996,000 | 1.5 (0.4-5.7) |  | 2.7 (0.6-11.5) |  |
| 4 | 27 | 5,275,000 | 1.0 |  | 1.0 |  |
| 5 | 147 | 5,430,000 | 6.2 (2.0-19.3) |  | 10.0 (2.7-36.7) |  |
| 6 | 112 | 6,312,000 | 5.3 (1.7-16.5) |  | 5.6 (1.6-20.0) |  |
| 7 | 52 | 5,131,000 | 2.8 (0.7-10.6) |  | 5.4 (1.2-23.6) |  |
| 8 | 91 | 5,629,000 | 3.3 (1.0-10.9) |  | 6.7 (1.7-26.3) |  |
| 9 | 93 | 6,924,000 | 4.7 (1.3-17.2) |  | 8.5 (2.0-35.6) |  |
| 10 | 64 | 4,702,000 | 3.5 (1.0-12.4) |  | 6.9 (1.6-29.2) |  |
| 11 | 57 | 4,561,000 | 4.6 (1.4-14.9) |  | 8.0 (2.1-31.0) |  |
| 12 | 72 | 5,084,000 | 3.2 (1.0-10.5) |  | 6.0 (1.6-23.1) |  |
| Gross provincial product* | - | - | 1.24 (0.96-1.59) | 0.10 | 1.16 (0.84-1.60) | 0.37 |
| Pig density* | - | - | 1.13 (0.88-1.46) | 0.34 | 0.93 (0.71-1.22) | 0.61 |
| Poultry density* | - | - | 1.11 (0.86-1.43) | 0.43 | 1.30 (0.95-1.78) | 0.10 |

- * Data were from 111 public referral hospitals in Thailand. All three continuous variables were standardized by centering at the mean values and divided by the standard deviation. The multivariable model consists of 809 patients with *Vibrio* spp. infection from 61,492,000 population over one year in 2022.

**Appendix A**

**STATA CODE FOR FITTING THE MULTIVARIABLE POISSON REGRESSION MODELS**

```
* Load the data
import excel using "TH_2022_74_provinces_NBD.xls" , sheet("Sheet1") firstrow

* Fit multilevel univariable Poisson regression model
global var "GPP_2021_S  Pig_density_S Poultry_density_S"
mepoisson Bps_PROV ib4.Health_region ///
 	, intmethod(mcaghermite) exposure(Pop_2022) ||Province_code:, irr
foreach var of global var{
mepoisson Bps_PROV `var’ ///
 	, intmethod(mcaghermite) exposure(Pop_2022) ||Province_code:, irr
}

* Fit multilevel multivariable Poisson regression model with province-specific random effects.
mepoisson Bps_PROV ib4.Health_region GPP_2021_S  Pig_density_S ///
 	Poultry_density_S,  intmethod(mcaghermite) exposure(Pop_2022) ///
||Province_code:, irr
	* Estimate p value for Health region variable
est store a
quietly mepoisson Bps_PROV          GPP_2021_S  Pig_density_S ///
 		Poultry_density_S,  intmethod(mcaghermite) exposure(Pop_2022) ///
||Province_code:, irr
	est store b
	lrtest a b
mepoisson Nts_PROV ib7.Health_region GPP_2021_S  Pig_density_S ///
 	Poultry_density_S,  intmethod(mcaghermite) exposure(Pop_2022) ///
	||Province_code:, irr
mepoisson Ssu_PROV ib11.Health_region GPP_2021_S  Pig_density_S ///
 	Poultry_density_S,  intmethod(mcaghermite) exposure(Pop_2022) ///
	||Province_code:, irr
mepoisson Vib_PROV ib4.Health_region GPP_2021_S  Pig_density_S ///
 	Poultry_density_S,  intmethod(mcaghermite) exposure(Pop_2022) ///
	||Province_code:, irr
```

**Appendix B**

**Examples of reporting criteria of relevant notifiable diseases reported to the national surveillance systems (NSS) in Thailand**

**Brucellosis**

Reporting suspected, probable and confirmed cases to Report506 (NSS in Thailand) using

ICD-10 codes: A23 (including A23.0, A23.1, A23.2, A23.3, A23.8 and A23.9)

- Suspected case is defined as a case with clinical criteria (at least one symptom) and a risk factor (e.g. drinking non-pasteurized milk or milk products or history of exposure to animals, animal tissues or secretions from animals)
- Probable case is defined as a case with clinical criteria, plus an epidemiological history associated with confirmed case or animals or a positive result according to presumptive diagnosis of laboratory criteria
- Confirmed case is defined as a case with clinical criteria plus a positive result according to specific diagnosis of laboratory criteria

Presumptive diagnosis of laboratory criteria includes

- Rose Bengal test (RBT) from serum is positive
- Lateral flow assay (LFA) from serum is positive

Specific diagnosis of laboratory criteria includes

- Pathogen identification based on bacterial culture or PCR
- Serology using Standard (Tube) agglutination test (SAT) using paired serum or ELISA or Coomb test from serum showing antibody level higher than that found using SAT 4-6 folds in case of acute infection and 16-256 folds in case of chronic infection
